# A Community-Based, Dietician-Led Nutrition Education Intervention to Improve Glycemic Control in Adults with Type 2 Diabetes in an Underserved Urban Community

**DOI:** 10.64898/2026.07.28.26359119

**Authors:** Reg Gorczynski, Samira Zarghami

## Abstract

**Background:** Diabetes is a major global public health challenge, associated with substantial medical, economic, and social burden worldwide. As clinicians working at the intersection of medicine and nutrition, we observed that despite advances in pharmacologic therapy, many individuals—particularly those living in underserved urban communities—continue to experience suboptimal diabetes control due to persistent barriers related to food insecurity, cultural food practices, and limited access to practical nutrition education. In response, we implemented a community-based pilot study in an underserved urban area that focused on dietitian-led nutrition education grounded in “Food Is Medicine” principles.

**Aims:** The intervention emphasized culturally responsive, cost-conscious, and client-centred approaches designed to make healthy eating simple and intuitive, with the goal of complementing standard diabetic medical therapy.

**Methods:** We used conventional diabetic marker responses, fasting blood sugar (FBS), HgbA1c (percentage of glycosylated hemoglobin, subsequently documented as A1c), and urinary albumin:creatinine ratio (UACR) in a dual cohort design study measuring whether groups receiving dietitian-led nutrition education in addition to conventional therapy would show improvements in diabetic markers relative to groups receiving conventional therapy alone.

**Summary:** Our pilot study summed over both cohorts support the hypothesis that additional dietitian-led nutrition education synergizes with conventional pharmaceutical therapy to improve diabetic control as gauged by improved FBS, A1c and UACR, implying a significant role for this combined approach to community based diabetic health care.

## Introduction

There have been substantial advances in the medical management of diabetes over recent decades, as reflected in successive updates to the Canadian Diabetes Association (CDA) clinical practice guidelines. These advances have expanded therapeutic options and improved glycemic control, as well as reduced the risk of long-term complications for many individuals with diabetes.

Despite these developments, lifestyle interventions—particularly diet and physical activity— remain foundational to both the prevention and management of type 2 diabetes. However, in routine clinical practice, time constraints often limit physicians’ ability to provide detailed lifestyle counselling. As a result, responsibility for nutrition education and self-management support frequently shifts to allied health professionals, including registered dietitians and diabetes nurse specialists, when such resources are available. Access remains inconsistent, and the cost of newer, highly effective pharmacologic therapies, such as sodium–glucose cotransporter-2 inhibitors (SGLT2i) and glucagon-like peptide-1 receptor agonists (GLP-1 RAs), continues to pose a significant barrier for many patients.

Emerging evidence suggests that the interaction between pharmacologic therapy and dietary intervention in diabetes management may be synergistic rather than merely additive, underscoring the importance of optimizing both approaches concurrently (Figure1). This concept has been highlighted in recent international scientific discourse and is supported by a growing body of literature demonstrating that targeted dietary patterns can enhance the effectiveness of pharmacologic treatments and improve glycemic outcomes [1–3].

**Legend to Figure 1.**
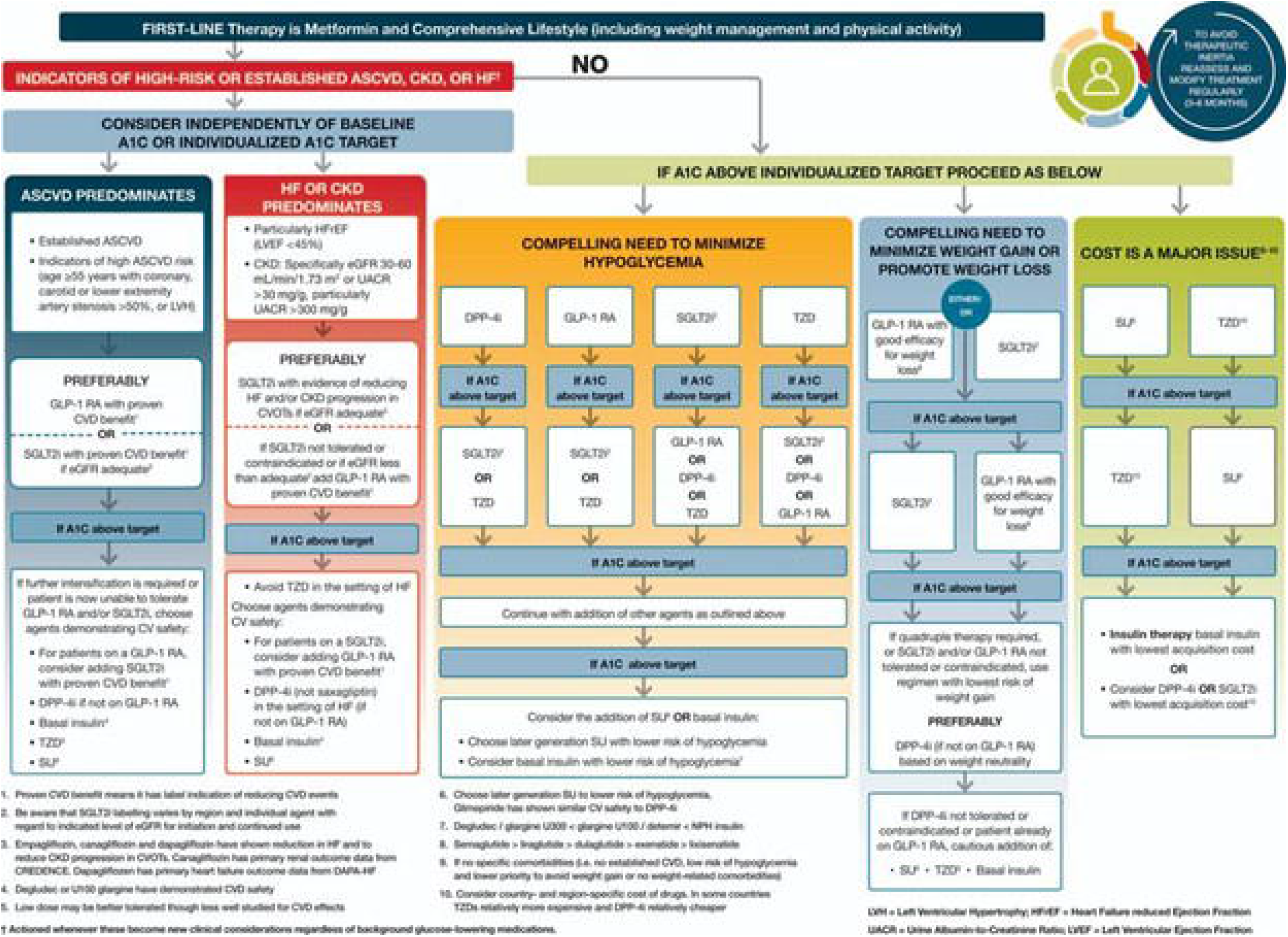
Taken from Diabetes Canada: Clinical Practice guidelines. Documents a stepped-care approach to diabetes management including pharmaceutical and lifestyle best practices.

Despite this wealth of information, in the absence of data evaluating the cost, effort, and time required for individuals to implement lifestyle changes compared with pharmacologic therapy, it remains challenging to advocate for the preferential selection of one approach over another (4-8). To address this gap, we designed a pilot, community-based study monitoring HbA1c and additional endpoints—including fasting blood glucose (FBS), serum creatinine/estimated glomerular filtration rate (Cr/eGFR), and urinary albumin-to-creatinine ratio (UACR)— in groups of volunteers in a two cohort design, to compare the relative effectiveness of additional structured lifestyle interventions versus pharmacologic therapy alone in improving diabetes management.

## Methodology

### Study Design and Setting

This was a prospective, community-based, randomized, delayed-intervention pilot study conducted over 16 weeks at a primary care/GP walk-in clinic serving an economically underserved urban population. The study protocol was approved locally, and all participants provided written informed consent prior to enrollment (Appendix 1).

### Participants

Participants were recruited from adult patients with non–insulin-dependent diabetes mellitus (NIDDM) routinely followed at the clinic. Eligibility was initially screened based on recent blood work showing HbA1c >7% and <8.5%. All participants received standard diabetic therapy including a sodium–glucose cotransporter-2 inhibitor (SGLT2i) for optimal diabetic treatment. No changes to diabetes medications were permitted during the study.

### Inclusion Criteria

Participants were asked to commit to a minimum of five clinic visits over the study period. Fasting blood work (FBW) was collected at baseline (week 0), week 8, and week 16, and body weight was recorded at each time point. There were no significant differences in baseline body weight between Group I and Group II (mean ± SD: 76 ± 15 kg). Participants were required to follow basic instructions for food purchasing and meal preparation. Any participants using continuous blood glucose monitoring (CBGM) were asked to log basal insulin use for future analysis; no changes in insulin use were reported during the study. Eligible participants were between 30 and 70 years of age, with no exclusions based on sex or ethnicity.

### Exclusion Criteria

Participants were excluded if they reported or demonstrated erratic adherence to NIDDM medications, either by self-report or documented clinical history, or if inconsistent adherence to dietary guidance was identified during follow-up appointments. Participants who changed diabetes medications during the study or expressed an inability or unwillingness to attend scheduled dietary education sessions were withdrawn. Where feasible, withdrawn participants were replaced with alternates drawn from the original recruitment pool; four such replacements were added to Group II.

### Dietary Education Intervention

The dietary intervention consisted of four structured group sessions delivered at two-week intervals by a registered dietitian and certified diabetes educator. Each 2-hour session combined interactive nutrition education with experiential learning and was conducted in community-accessible settings. Nutrition education was personalized, client-centered, and culturally responsive, grounded in the **Food Is Medicine framework**, and delivered using a dietitian-developed approach designed to simplify healthy eating concepts and enhance comprehension, visualization, and practical application. Sessions emphasized making healthy eating simple and intuitive, integrating participants’ traditional foods, and supporting **glycemic control, gut–brain health, gut microbiome balance, and cognitive well-being**.

Experiential supermarket tours complemented classroom sessions by addressing real-world challenges related to food access, affordability, and decision-making. Participants learned to identify nutrient-dense, cost-conscious options, compare food labels, understand fiber content and carbohydrate quality, and reduce reliance on ultra-processed foods. The supermarket environment also provided a sensory and visual learning space that minimized language barriers and facilitated discussion of culturally specific foods and cooking practices. Rather than prescribing a specific diet, participants were supported in adapting familiar meals to align with evidence-based nutrition principles [4-6].

### Randomization and Group Allocation

From an initial pool of 26 eligible participants, 20 were randomly assigned to one of two groups (n = 10 per group). Group I received the dietary intervention during weeks 0–8, while Group II continued usual care and served as a control. Six additional participants were retained as alternates to replace dropouts. At week 8, Group II crossed over to receive the same dietary intervention during weeks 8–16, while Group I returned to usual care. At crossover, Group II participants had not previously received any dietary intervention and thus constituted a naïve cohort at initiation of the intervention (see schematic)

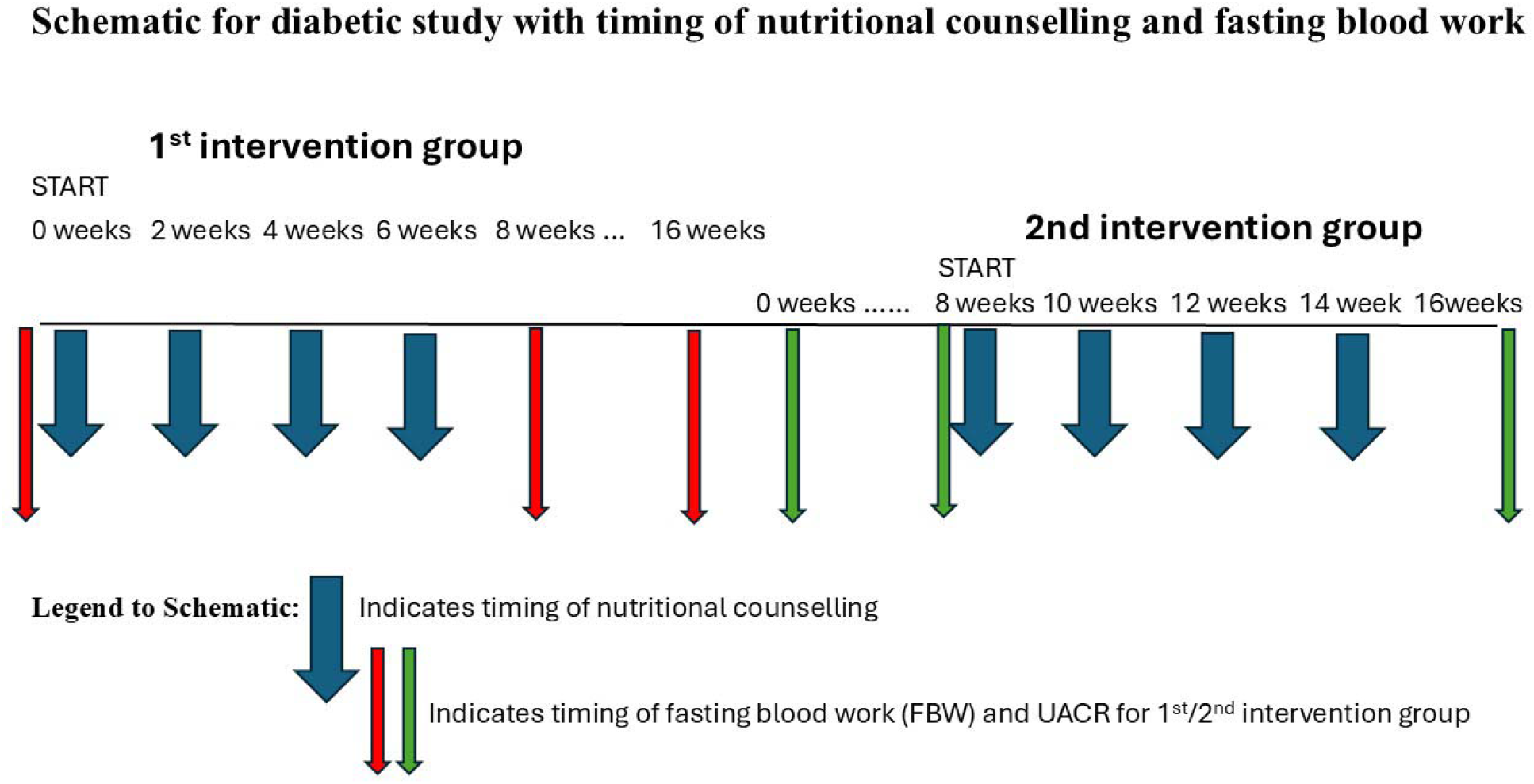

### Outcome Measures

Fasting blood work (FBW), urinalysis, and body weight were obtained at baseline, week 8, and week 16. Primary outcomes included HbA1c, fasting blood glucose (FBS), and urinary albumin-to-creatinine ratio (UACR). Secondary laboratory measures included serum creatinine, estimated glomerular filtration rate (eGFR), electrolytes, and serum albumin.

### Statistical Analysis

Data were analyzed using statistical methods described in the Figure Legends. Between-group comparisons of HbA1c, FBS, UACR, and body weight were conducted at weeks 0, 8, and 16, and within-group changes over time were also assessed. Results are presented as mean ± standard deviation with respect to controls with statistical probability (t-test) recorded as p<0.05 (see Figure 2 below). In a final analysis data for all subjects in the two groups were pooled at initiation/completion (8 weeks) of dietary intervention and compared (ANOVA) using a Wilcoxon Rank Sum analysis, again with statistical probability recorded as p<0.05 (see Figure 3 below). Post Hoc G-power analysis are recorded in the Figure legends for a 1-tailed test comparing two groups.

**Legend to Figure 2.**
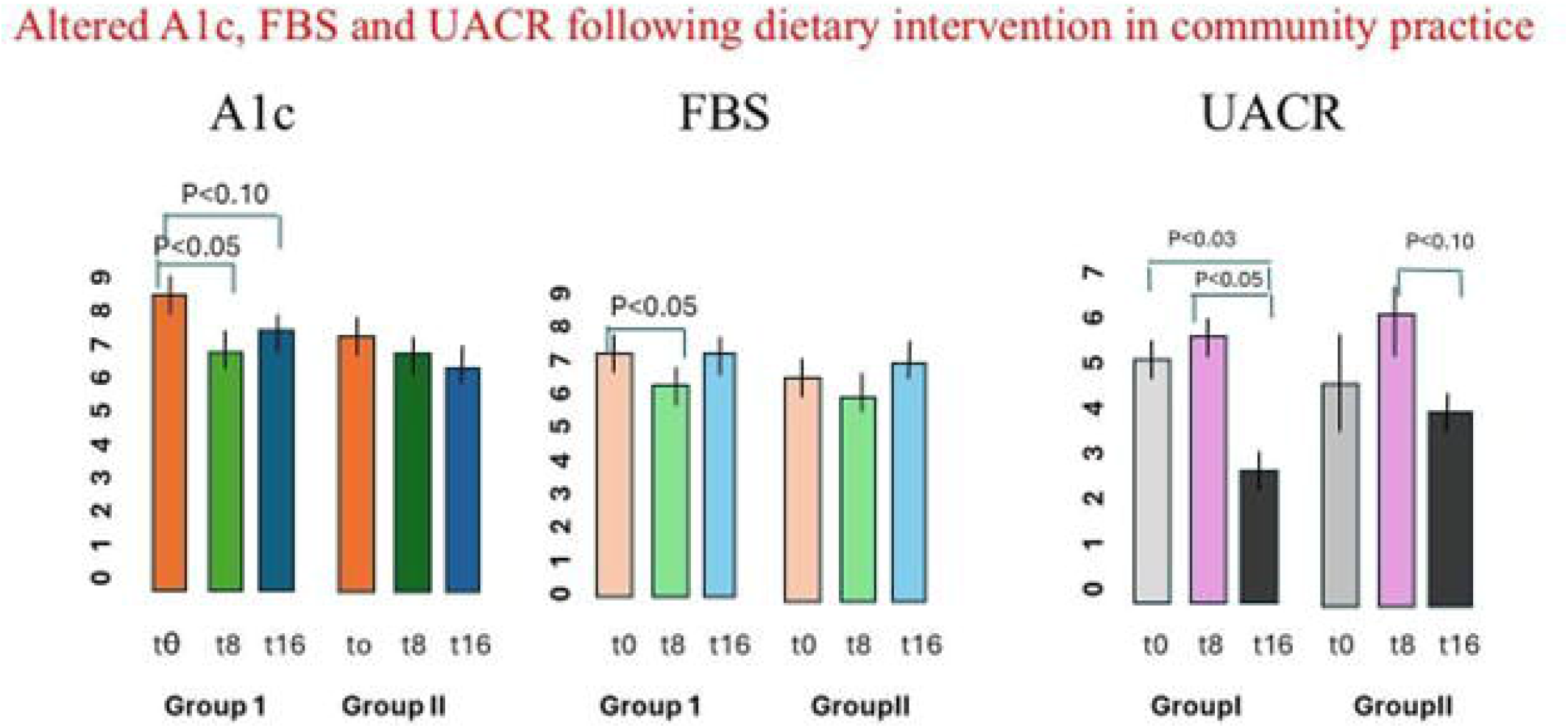
Mean (within groups) of A1c, FBS or UACR at initiation (week 0) and at 8 and 16 weeks of study. Note that Group I received dietary instruction only from weeks 0-6, while group II only from weeks 8-14. Data represent arithmetic mean±SD. P values used Wilcoxon rank sum tests for comparison.

**Legend to Figure 3.**
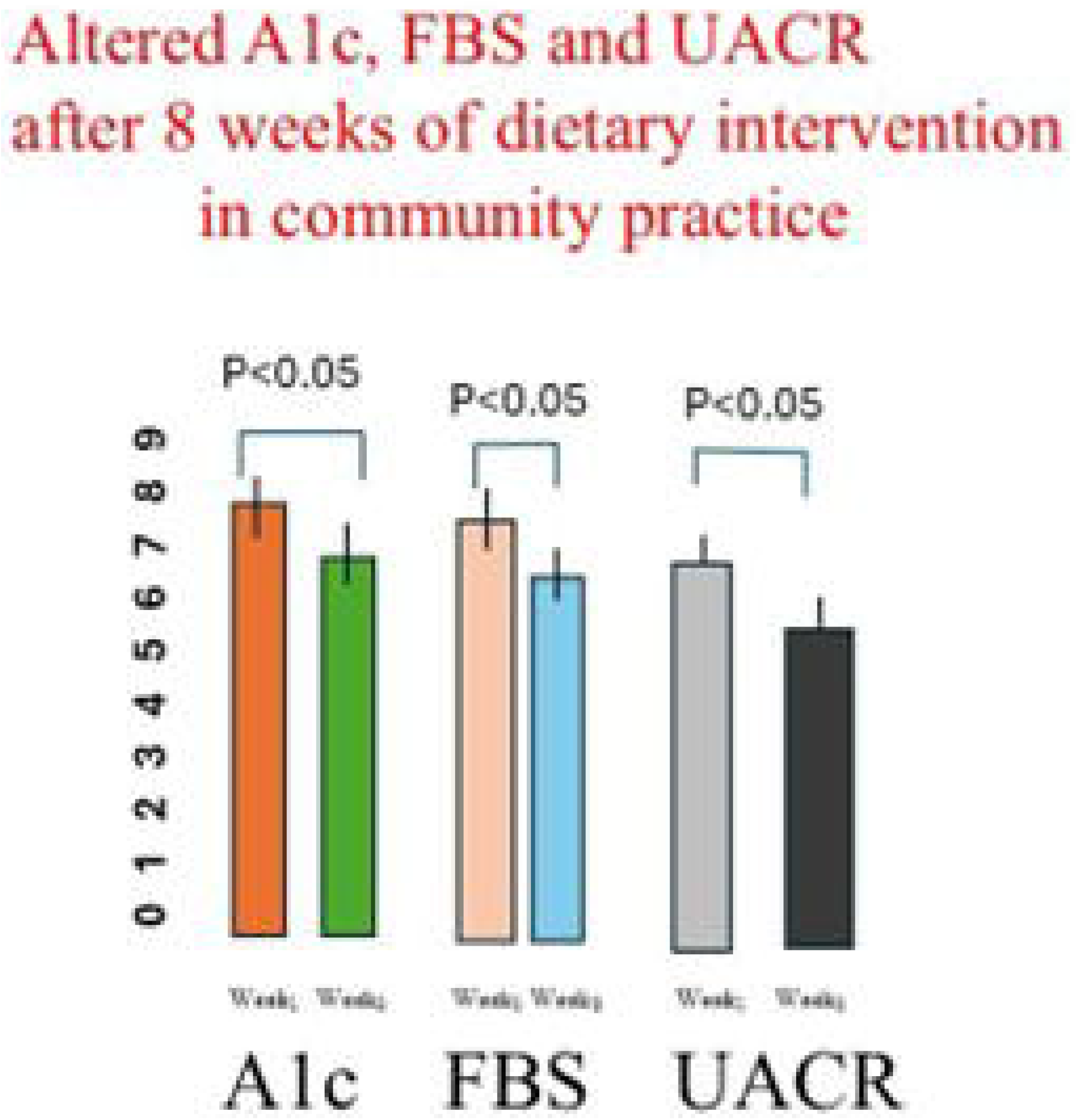
Comparison of A1c, FBS and UACR in pooled groups of comparable subjects at baseline, and after 8 weeks of nutritional instruction from a registered dietician (see text and Legend to Figure 1 for more details). While 20 subjects started the study, only data for 17 subjects were available for the 8 week follow-up measures, following “drop-outs” after non-adherence to the study design. Data represent mean ±SD, with p values determined by Wilcoxon Rank Sum.

## Results

As indicated in the Methodology section, we focused our assessment on data obtained from Fasting blood work (FBW), urinalysis, and body weight measured at baseline, week 8, and week 16 in 2 groups of 10 subjects (Ss) per group, receiving dietary instruction (see schematic above) at over different time schedules. In the first analysis shown in Figure1, we have focused for the sake of simplicity on the following primary outcomes: HbA1c, fasting blood glucose (FBS), and urinary albumin-to-creatinine ratio (UACR). We recovered additional data also on secondary laboratory measures, which included serum albumin, sodium (Na□) and potassium (K□), serum creatinine (Cr), estimated glomerular filtration rate (eGFR), and the Cr:eGFR ratio. In all of these latter cases, however, no significant changes were observed within or between Groups I (1^st^ intervention group) and II (2^nd^ intervention group)-data not shown. Only a possible trend (p∼0.1) was seen for (Cr, eGFR and Cr:eGFR ratio), so none of these data are shown to maintain clarity and focus on the key outcomes.

All data subsequently shown in Figures 2 and 3 focus then on group differences in A1c, FBS and UACR. There are six panels Figure1 (read left to right). The first two panels present serum A1c measures over time. In Group I, there was an improvement in diabetes control following dietary instruction, as indicated by the decline in A1c after the 8-week dietary intervention (first two bars), an improvement which showed a trend to persist through week 16, during which no additional dietary intervention was provided (3^rd^ bar of this group). In contrast, in Group II (with no dietary intervention in the first 8 weeks of study) no change in A1c between weeks 0 and 8 (4^th^ and 5^th^ bars in this first panel), serving as a control for Group I during this period. Between weeks 8 and 16, however, when group IIM did receive dietary advice, there was only a statistically insignificant change in A1c, with a downward trend observed (p ∼ 0.1).

It is important to note that 9 of 10 participants in Group I completed the study through to week 16 (1 Ss lost between weeks 8 and 16). In Group II, however, four participants initially assigned to the group had dropped out prior to week 8, and two additional participants (from the “new” Group II, which included both original and replacement participants) failed to complete the final 8 weeks, leaving complete data for only 8 of 10 participants. We hypothesize that the lack of significant A1c change in Group II between weeks 8 and 16 reflects technical limitations rather than a true absence of a reproducible effect of dietary intervention. [In fact, when data were later pooled across groups just with regard to pre/post dietary intervention (Group I: weeks 0–8; Group II: weeks 8–16), a significant decline was observed in 17 of 20 participants (p < 0.05; see Figure 3 below)].

Comparison of the next six panels in Figure 2, studying changes observed in FBS as a measured variable to monitor diabetes control, is revealing and provides additional support for the trends observed in the first six panels. There was again a significant decline in Group I between weeks 0 and 8, consistent with the reductions seen in A1c, although this decrease in FBS did not persist to week 16 in the absence of further diet instruction. In contrast, FBS in Group II showed no significant change at any time from weeks 0 through 16. As mentioned above with regards to A1c levels, the lack of change in FBS in Group II during the first 8 weeks serves as a useful control for the validity of diet intervention for the improvement observed in Group I. Again we hypothesize that the absence of a comparable change in Group II between weeks 8 and 16 may be attributable to technical factors associated with subject “drop-out” rather than a true absence of reproducibility of the diet intervention effect. [Once again we have attempted to address this issue by pooling comparable data across groups (Group I: weeks 0–8; Group II: weeks 8–16), to show a significant decline in FBS after 8 weeks of dietary intervention, now averaged over all 17 participating Ss.-see Figure 3 below].

The final series of panels in Figure 2 examines group difference in urinary albumin-to-creatinine ratio (UACR). Interestingly, now in Group I there was no significant change observed by week 8, at the end of dietary intervention, but by week 16, a significant decline in UACR was noted relative to both baseline and week 8, even though no further dietary intervention was made for this group from weeks 8 to 16 (and changes in A1c and FBS declined). We hypothesize that dietary intervention can indeed induce meaningful improvements in renal function, but these may manifest more slowly than changes in A1c or FBS. This might help explain a trend towards lower UACR that was observed in Group II by week 16.

The data above, for these 3 outcome measures (A1c, FBS and UACR) provide some support for the idea that a short-term dietary intervention (8 weeks) could improve diabetes control. Limitation in group size (10 Ss was felt to be the maximum which could receive the individual dietary instruction we had planned), and also the thought that a cross-over design would allow a better control design to assess the hypothesis regarding an important role for dietary intervention, contributed to significant flaws in the study outcome. In particular, this design we hypothesize contributed to the major drop-off in participation in group II Ss, who had been recruited but essentially “left alone” to usual care for 8 weeks-as noted earlier 9/10 Group I Ss completed all 16 weeks of study, while even with 6 replacements for drop-outs (3 at each time point), only 8 Ss remained in group II at 16 weeks. We thus asked the question whether our data would show better reliability in assessing the value of dietary intervention itself by simply comparing pre-/post diet intervention for all Ss after simply 8 weeks of such intervention. Pooling data regardless of whether diet intervention took place between weeks 1 and 8 (group I) or 8 and 16 (group II), we compared all 17 subjects from pre-/post diet intervention, with data now shown in Figure3.

These data re quite striking. They indicate quite robustly that for all, the measures used, the pooled data across both groups indicates a very significant decline in A1c, FBS and in UACR, after 8 weeks of intervention (Figure 2). We suggest this pooled data supports the hypothesis that the dietary interevent improved diabetic control in these Ss, and furthermore that there was a concomitant impact of dietary education on renal health. Once again (data not shown; P<0.1) even with these pooled data no significant differences were seen nevertheless in other measures (serum albumin, sodium (Na□) and potassium (K□), serum creatinine (Cr), estimated glomerular filtration rate (eGFR), and the Cr:eGFR ratio).

## Summary and Conclusions

There is increasing interest in understanding how structured dietary interventions can impact objective measures of diabetes management, including blood glucose control, A1c levels, and renal function. Comprehensive nutrition education has been highlighted as a critical strategy to empower patients to choose foods that are healthy, affordable, culturally relevant, and easy to prepare, supporting better management of diet-related chronic diseases such as type 2 diabetes, high blood pressure, and cardiovascular disease [7–9].

Our findings from this small community-based study support these ideas. In a randomized controlled pilot trial, dietary intervention produced significant and sustained improvements in fasting blood glucose, A1c, and renal function as measured by UACR over 8–16 weeks. Importantly, these outcomes highlight the effectiveness of making healthy eating simple and intuitive, grounded in the “**Food Is Medicine”** framework. By embedding culturally sensitive, client-centered education that incorporates participants’ traditional foods, addresses food cost, and facilitates practical meal planning, participants were empowered to make informed, real-world food choices. This approach appeared to promote intuitive healthy eating behaviors, resulting in improved glycemic control, gut–brain health, and overall metabolic function, ultimately enhancing diabetes management. Health literacy is now documented to be an important component of glycemic control and nutritional status in diabetic individuals [10].

We acknowledge that a limitation of the study was the relatively small number of participants who completed all sessions. Randomization at initiation placed one cohort in a delayed-engagement group, which made it challenging to maintain full participation in the second cohort. As a result, data from this group, did not fully replicate findings from the initial cohort. As an example, trends in decline in UACR in Group I seen at week 16 might suggest that physiological and metabolic improvements observed in this first group might have begun to emerge in the second cohort, even though follow-up was shorter for these participants (8 weeks) compared with the first intervention group (16 weeks). Indeed the pooled comparable date for the 2 groups shown in Figure 3 further support the observation that changes in A1c, FBS, and UACR can occur within just 8 weeks of dietary intervention.

Future pilot studies aiming to replicate our findings should aim to engage larger cohorts (≥15 per group, from an initial volunteer pool of ∼40) and implement strategies to maintain participant engagement throughout the study period, including for cohorts not actively receiving dietary instruction. Finally, given that UACR is known to be sensitive to blood pressure changes—and some 90% of participants were on antihypertensive medications—future studies should monitor blood pressure to account for this potential confounder.

Overall, our results reinforce the concept of a **dual approach to diabetes management**, highlighting the synergistic potential of **client-centered, culturally responsive dietary education** and pharmacological treatment. We have demonstrated that nutrition education framed around the “**Food Is Medicine”** concept, and practical strategies for making healthy eating simple, can lead to intuitive healthy eating behaviors and measurable improvements in glycemic and renal outcomes. Our study further shows that such interventions can reduce healthcare costs, improve chronic disease management, and promote long-term health. Furthermore, these findings highlight the importance of incorporating structured nutrition education alongside pharmacological strategies in routine diabetes care to optimize patient outcomes. This can be done even in scenarios where patients opt to use food delivery services [11]. More sophisticated analysis suggests even further benefits from attention to nutritional status on the overall health (including immunological health) of diabetic patients [12].

As an addendum, we note that without exception all study participants highly valued the sessions. They consistently described the culturally relevant and simplified nutrition education as practical, impactful, and empowering, noting that it strengthened their confidence and improved their ability to influence their own diabetes control beyond that attributable to medications alone. We believe further support for the relevance of our study to diabetes care can be seen in a new publication by Christensen et al (13). This highlights some concerns re the possible nutritional and overall health consequences of the “rush” to focus on introduction of novel diabetic medications (GLP-1RA etc.) before a thorough exploration of their widespread use in ethnically, culturally and financially diverse populations, and without simultaneously addressing those consequences.

## Data Availability

All data produced in the present study are available upon reasonable request to the authors

## Acknowledgements

The authors would like to thank management (Dr. Jauher Ahmed and Cidalia Ganesh) and all of the staff at the Intrepid Clinic (Jane-Finch) for their help/support, without whom this study would not have been possible.

## Notes

**Funding** This study was supported entirely from local funds within the medical clinic, and by personal financing for publication by RMG.

**Conflict of Interest:** Both authors confirm there are no conflicts of interest to declare

### Competing Interest Statement

The authors have declared no competing interest.

### Author Declarations

Ethics Committee of Intrepid Health gave full ethical approval for this work.

