## Supplemental File#1 for "A Community-Based, Dietician-Led Nutrition Education Intervention to Improve Glycemic Control in Adults with Type 2 Diabetes in an Underserved Urban Community"

**APPENDIX 1: Informed consent**

I, XXXXX , have read the brief outline of a proposed study to be conducted by XXXX and XXXXX at the XXXX Clinic beginning in early 2025, entitled "**A NIDDM community study (8 weeks) on dietary intervention and diabetes management"**

1. I understand that Diabetes is a major global chronic disease issue, afflicting >10% of the world population, and causing significant medical, economic and social emotional cost world-wide. I understand that there are many old and new drugs which have improved the ability to monitor and control blood sugars, and also the late developing complications of diabetes, including cardiovascular disease, renal failure, and ophthalmologic disorders. I also understand that there is a recognition that diet advice and weight control is known to be an important component of optimal diabetes management. Given the costs of payment for drugs/dietary needs, particularly in the current economic climate, I understand the need to be able to have an appreciation of a comparison of how much of an influence drugs vs diet have on blood sugar control. I understand that this study is designed to try to obtain information which will provide information on this comparison. I confirm that I have **NOT** been told that this study will definitely improve my blood sugar management, and that I know I have to continue to take my usual diabetic medications throughout the study. I must report any failure to take those medications, and realize that doing so ***will exclude me*** from further participation in the study.

2. I understand that I am being offered the opportunity to engage in this study, which will involve my completion of 4 independent clinic group sessions along with another 11 individuals. Each session will involve an approximately 2hr meeting with a Registered Dietitian to discuss optimizing food purchases and meal planning/preparation which fit within my ethnic restrictions. These sessions will take place in a community meeting place (TBD) and in some cases with kitchen access. Where cooking is a component of the session I will provide at least some (if not all) of the materials I routinely use for cooking at home.

I understand that there will be two study groups each independently completing the 8-week study, and I **may *not*** be enrolled in the study until after the first group (at 8 weeks) has completed their diet education.

3. I understand that I will have urine and weight testing, as well as routine fasting blood work drawn, at initiation of the study, and after 8/16 weeks of the study. On completion of the study I will receive a VISA gift card for $100 for attendance at all the educational sessions and blood work donation.

4. Within 16 weeks of study completion I understand I will be invited to a “debriefing” session with XXX and XXX, where I will be able to provide my own thoughts on what I liked/valued or disliked about the study, and where I will hear at least preliminary results from this study.

5. I hereby give my consent/authorization to XXX and XXX to publish the results of their study to as wide an audience as they feel are appropriate to hear this information, with the restriction that there will be **NO** identification of any participants, who shall always remain anonymous.

**SIGNED** **DATE:**
